# Pre-treatment biopsychosocial predictors of chemotherapy-induced peripheral neuropathy trajectories in people with breast cancer

**DOI:** 10.64898/2026.05.13.26353023

**Authors:** Charles-Antoine Auger, Antoine Frasie, Maud Bouffard, Frédérique Therrien, Sarah Béland, Anne Dionne, Robert H. Dworkin, Lucia Gagliese, Jennifer S. Gewandter, Philip L. Jackson, Sophie Lauzier, Julie Lemieux, Josée Savard, Lynn R. Gauthier

## Abstract

**Purpose:** Chemotherapy-induced peripheral neuropathy (CIPN) affects many people receiving taxane treatment for breast cancer. Symptom trajectories vary, with some recovering, and others experiencing persistent, or delayed worsening (coasting) symptoms. The prevalence and predictors of these trajectories remain unclear. This study identified the prevalence and biopsychosocial predictors of CIPN persistence, improvement, and coasting within three months post-treatment.

**Methods:** This secondary analysis included participants treated with taxanes for stage I-III breast cancer who completed the Functional Assessment of Cancer Therapy/Gynecologic Oncology Group–Neurotoxicity-4 (FACT/GOG-NTX-4) at baseline, post-chemotherapy, and three months later. A minimally important difference (MID) from baseline on the FACT/GOG-NTX-4 defined persistence, improvement, coasting, and no MID-CIPN (below the MID threshold at each assessment) trajectories. Baseline assessments included self-reported pain/well-being, sensory, balance, and lower limb physical functioning measures, and sociodemographic and treatment data were collected.

**Results:** Among 102 participants (51.57±11.24 years), persistence occurred in 34.3%, improvement in 25.5%, coasting in 6.9%, and no MID-CIPN in 33.3%. Compared to no MID-CIPN, older age (OR=1.120; 95%CI: 1.026–1.222), higher expected pain (OR=1.630; 95%CI: 1.082–2.456), and cold hyperalgesia at the foot (OR=1.130; 95%CI: 1.018–1.254) predicted persistence. Lower fatigue predicted improvement (OR=0.904; 95%CI: 0.845–0.968). No predictors were identified for coasting.

**Conclusion:** CIPN trajectories are heterogeneous. Age and pre-treatment pain expectations, cold hyperalgesia, and fatigue differentiate patients with persistent CIPN and those likely to improve from those with no CIPN.

**Implications for Cancer Survivors:** Early identification of individuals at risk for persistent neurotoxicity may support risk stratification and guide targeted supportive care strategies.

## Introduction

Chemotherapy-induced peripheral neuropathy (CIPN) is a common and disabling effect of neurotoxic chemotherapeutic agents [1]. Symptoms can include tingling, numbness, reduced touch sensitivity, pain, muscle weakness, and balance difficulties, primarily affecting the distal extremities (hands and feet) [2, 3]. These symptoms impair quality of life (QOL) and are associated with anxiety, depression, and psychological distress [4, 5]. Despite its burden, preventive or curative treatments are not available, and symptom management remains limited [1].

In people receiving taxane-based chemotherapy for breast cancer, the prevalence of CIPN at treatment completion ranges from 43% to 74% [6, 7]. CIPN symptoms are reported to typically develop within the first two months of chemotherapy, progress during active treatment, and stabilize shortly after treatment completion [8]. However, symptom resolution post-treatment is variable [2]. In addition, some people experience a delayed onset or worsening of CIPN symptoms after completion of chemotherapy, an effect known as coasting. While coasting has been clinically described, its frequency remains poorly quantified [9, 10].

Among taxane-treated breast cancer survivors, CIPN prevalence one to three years post-diagnosis varies widely, from 11% to over 80% [11]. This wide variation in prevalence estimates may reflect not only methodological differences in study design, follow-up duration and timing, assessment methods, and differences in regimens of neurotoxic chemotherapies [12, 13], but also heterogeneous symptom trajectories, including persistence, improvement, and coasting.

Trajectory-based research (i.e., the patterns by which symptoms emerge, persist, or resolve over time) has increasingly been used to better understand variability in cancer-related symptoms, including fatigue, depression, and pain [14–19]. These studies demonstrate heterogeneous symptom trajectories that may be associated with multidimensional factors including age, comorbidities, socioeconomic disadvantage, pre-and acute post-treatment symptom burden, and psychosocial factors [14–19]. In the context of breast cancer–related pain, two trajectories in the first year post-diagnosis have been identified: a persistent trajectory (56%) and a transient trajectory (44%) [14]. Patients in the persistent trajectory group reported greater pain intensity at 6 and 12 months than patients in the transient pain trajectory group. In addition, greater baseline psychosocial vulnerability, including cognitive, emotional, social, and QOL factors, predicted persistent pain trajectory membership [14].

While trajectory-based approaches have begun to elucidate the complexity of the cancer symptom experience, our understanding of CIPN trajectories remains extremely limited. To date, only one study has applied a trajectory-based approach to CIPN [20]. This study identified two trajectories among 75 people treated with vincristine for lymphoma: a high CIPN trajectory (41%) and a low CIPN trajectory (59%). The high CIPN trajectory was characterized by increasing CIPN severity during chemotherapy, peaking at treatment completion, and decreasing slightly at ten weeks post-treatment. The low CIPN trajectory was characterized by low CIPN severity during treatment that gradually decreased at ten weeks post-treatment. Compared to people in the low CIPN trajectory, people in the high CIPN trajectory were older and less likely to be employed. While this study provides important data documenting heterogeneous CIPN symptom courses, the restricted sample size (n = 75) may have limited the range of detectable trajectories, as group-based trajectory modeling typically requires substantially larger samples to reliably identify distinct subgroups [21], potentially oversimplifying symptom evolution. As a result, some trajectory patterns may not have been fully captured. Moreover, CIPN trajectories have not been examined in people treated with taxane-based chemotherapy for breast cancer.

CIPN risk factors span biopsychosocial domains. Older age, higher body mass index (BMI), baseline neuropathy, cumulative chemotherapy dose, medical comorbidities, anxiety, depression, and fatigue have been associated with CIPN in people with breast cancer [22–26]. However, these studies have primarily focused on predicting overall CIPN incidence or severity measured at discrete time points, rather than on how risk factors relate to distinct CIPN trajectories, thereby limiting risk stratification and tailored monitoring and intervention strategies. Accordingly, the present study aims to (1) determine the prevalence of CIPN persistence, improvement, and coasting trajectories in the first three months following chemotherapy in people receiving taxane-based treatment for breast cancer, and (2) identify pre-treatment biopsychosocial predictors of these trajectories.

## Methods

### Study Design, Setting and Participants

This secondary analysis was conducted using data from a prospective longitudinal study of CIPN measurement carried out between 2017 and 2019. As previously described [27], people with early-stage breast cancer (stage < IV) were recruited prior to the initiation of taxane-based chemotherapy at the Centre des maladies du sein, CHU de Québec–Université Laval (Quebec, Canada). Eligible participants were aged ≥ 18 years, chemotherapy naïve, and able to read, write, and understand French. Exclusion criteria included the presence of metastatic disease, comorbidities affecting peripheral sensory function (e.g., HIV/AIDS, diabetes, alcohol use disorder as defined by NIH criteria [28], or polyneuropathy), and scores < 20 on the Short Orientation Memory Concentration Test [29], a brief measure of cognitive impairment that has been used in other studies involving people with cancer [30, 31]. The study was approved by the Research Ethics Committee of the CHU de Québec-Université Laval (REB#2017-3312). It is reported in accordance with the Strengthening the reporting of observational studies in epidemiology (STROBE) guidelines for cohort studies (see Supplementary Table S1 for the STROBE checklist).

Written informed consent to participate was obtained prior to data collection and included authorization to access participants’ electronic medical records (EMR). Study visits were scheduled before the first chemotherapy cycle or within two weeks for regimens not starting with a taxane (T0). The second assessment (T1) was conducted at the time of the final taxane infusion (±1 week). The third assessment (T2) took place three months after the final taxane infusion.

At T0, sociodemographic, cancer, treatment, and clinical data were collected from a brief participant interview and the EMR at baseline, and participants completed self-report measures assessing pain, fatigue and sleep, psychosocial wellbeing, QOL, a brief Quantitative Sensory Testing (QST) protocol [32], and a lower limb physical functioning assessment. To minimize order effects, the order of questionnaire completion, QST, and physical functioning assessments at T0 was randomized.

### CIPN trajectory groups

At T0, T1, and T2, participants completed the Functional Assessment of Cancer Therapy – Taxane (FACT-Taxane) [33]. To determine trajectory group membership, the Functional Assessment of Cancer Therapy/Gynecologic Oncology Group – Neurotoxicity 4-item scale (FACT/GOG-NTX-4) [34] was calculated from the first four items of the FACT-Taxane, assessing numbness, tingling, and discomfort in the hands and feet on a 5-point Likert scale ranging from 0 (“*not at all*”) to 4 (“*very much*”). Items are reverse-scored and summed to a total score ranging from 0 to 16, with higher scores reflecting lower neurotoxicity, in accordance with the scoring instructions of the instrument [34]. The FACT/GOG-NTX-4 has been used as an outcome measure in observational studies [35, 36] and has shown strong correlations with the Common Terminology Criteria for Adverse Events (CTCAE) grades (*r* = 0.67 – 0.80) [37].

CIPN trajectory groups were determined using a clinical anchor-based method to establish trajectory group membership [37]. Following the methods of Li et al. [37], using FACT/GOG-NTX-4 scores calculated according to standard instructions, the sign of change scores was reversed prior to threshold application, such that a positive change indicates worsening neurotoxicity. Therefore, a change score of ≥ 3.39 points on the FACT/GOG-NTX-4 from T0 was considered a minimally important difference (MID) in sensory symptoms, which correspond to a shift from CTCAE grade 0 (“*no symptoms*”) to grade 1 (“*paresthesia not interfering with function*”) [37]. Although the MID is typically used to capture change between two time points, in this study it was applied as a threshold at each follow-up assessment (T1 and T2) to determine whether the increase in CIPN symptoms was at least ≥ 3.39 points relative to T0 (i.e., meeting criteria for MID-CIPN). Depending on whether the MID threshold was met at T1 and/or T2, participants were classified into one of four trajectory groups: Persistence (MID-CIPN at both T1 and T2), Improvement (MID-CIPN at T1 only), Coasting (MID-CIPN at T2 only), and No CIPN (no MID-CIPN at either T1 or T2).

### Candidate predictors: Sociodemographic, clinical, and treatment characteristics measures

As previously reported [27], clinical data, including cancer and treatment characteristics, medical comorbidities, and prescribed medications were extracted from the EMR and the CHU de Québec-Université Laval oncology registry. A brief interview was conducted to collect sociodemographic information and pain history. The Charlson Comorbidity Index (CCI) [38, 39] and the Anticholinergic Drug Scale (ADS) [39] were also calculated by the research assistant (RA).

### Candidate predictors: Self-report measures of pain, fatigue and sleep, psychosocial wellbeing, and quality of life

Pain presence related to cancer and other causes over the past three months was assessed using the Pain History Questionnaire (PHQ) [40]. At T0, participants also reported how much pain they expected to have immediately after (“How much pain do you expect to feel immediately after your chemotherapy?”) and one week after chemotherapy (“How much pain do you expect to feel one week after your chemotherapy?”) using single-item numeric rating scales ranging from 0 (*no pain*) to 10 (*worst possible pain*). The mean of these two items was calculated to derive an expected pain variable [41]. Pain severity and interference were measured with the Brief Pain Inventory (BPI) [42], which includes four items assessing pain intensity and seven items assessing pain-related interference with daily functioning, all rated from 0 (*no pain/no interference*) to 10 (*worst pain/worst interference*). Non-neuropathic, neuropathic, and affective pain qualities were assessed with the Short-Form McGill Pain Questionnaire-2 (SF-MPQ-2) [43], a 22-item measure in which each item is rated from 0 (*no*ne) to 10 (*worst possible*). Neuropathic pain symptoms, including spontaneous, paroxysmal, and evoked pain, along with dysesthesia and paresthesia, were measured using the Neuropathic Pain Symptom Inventory (NPSI) [44], a 10-item questionnaire in which each symptom is rated from 0 (*no symptom*) to 10 (*worst symptom intensity*).

Sleep quality and cancer-related fatigue were assessed using the Pittsburgh Sleep Quality Index (PSQI) [45] and the Functional Assessment of Chronic Illness Therapy (FACIT)-Fatigue Scale [46]. Cancer-related QOL was measured with the four subscales of the FACT-General [47]: physical well-being (PWB), social/family well-being (SWB), emotional well-being (EWB), and functional well-being (FWB). Depressive symptoms were evaluated using the total score of the Center for Epidemiologic Studies Depression Scale (CES-D) [48], and negative pain appraisals were evaluated with the total score of the Pain Catastrophizing Scale (PCS) [49]. All instruments have been validated for use with people with cancer or chronic non-cancer pain. Higher scores on the FACIT-Fatigue and on the four subscales of the FACT-General (PWB, SWB, EWB, FWB) indicate lower fatigue and better QOL. Higher scores on the other measures reflect greater pain, symptom burden, and psychological wellbeing.

### Candidate predictors: Thermal, vibration, and mechanical detection threshold measures

QST was conducted by a trained RA using an adapted protocol [32]. Assessments included cold (CPT) and heat pain thresholds (HPT), as well as cool (CDT), warm (WDT), vibration (VDT), and mechanical detection thresholds (MDT). Thermal and vibration thresholds were measured on the dorsum of the right hand and foot [39, 50] with the TSAII NeuroSensory Analyzer and the VSA 3000 (Medoc, Israel), and MDT was evaluated using the SenseLab Aesthesiometer II (Somedic, Sweden). Thermal thresholds were assessed using a 3×3 cm thermode that delivered thermal stimuli, starting at 32 °C with a change rate of 0.5 °C/sec and capped at 0 °C (cooling) and 50.5 °C (warming). The temperature at first report of sensation and pain and the time, in seconds, from stimulation onset to patient report via mouse click, were recorded. VDT testing began at 0 µm/sec and increased at a rate of 1 µm/sec. Participants were instructed to indicate the moment they first perceived the vibration. MDT was assessed using hand-held nylon filaments applied in ascending order of force. Once the stimulus was perceived, testing continued in descending order using the next larger filament. The smallest filament diameter that evoked a touch sensation was recorded. Threshold was calculated as the geometric mean of six filament forces and the mean of three thermal and vibratory trials per test site. Higher WDT, HPT, VDT, and MDT values indicate lower sensitivity, whereas higher CDT and CPT values indicate greater sensitivity.

### Candidate predictors: Performance-based outcome measures of physical functioning and observer-rated functional status

The Timed Up and Go (TUG) test [51], averaged over two trials, was used to evaluate balance and mobility. Balance was measured using the Short Physical Performance Battery (SPPB) [52] and information on walking aid use and the number of falls or near-falls in the past week was collected by the RA. Functional status was assessed by the RA using the Karnofsky Performance Status Scale (KPS) [53], an observer-rated scale used to assess functional status.

### Statistical analyses

The sample size was determined by the number of participants with complete data on the FACT/GOG-NTX-4 at T0, T1, and T2 [27]. A post hoc sensitivity power analysis was conducted using G*Power 3.1. The study had an 80% power to detect a minimum effect size of f² = 0.18 (R² ≈ 0.15) in the regression analysis, corresponding to a medium effect [54]. All analyses were performed using IBM SPSS Statistics for Windows, version 30. Data were cleaned and checked for missing values. For scales with ≤20% missing items, missing values were imputed using the participant’s mean item score within the corresponding scale [55]. Otherwise, the specific scale score was treated as missing. Analyses were conducted using the imputed dataset. Descriptive statistics were used to characterize participants’ sociodemographic characteristics, disease and treatment factors, and study-related measures, as well as the prevalence of each trajectory group.

To determine candidate predictors for inclusion in the regression model assessing pre-treatment predictors of CIPN trajectories, bivariate analyses were first carried out between the candidate predictors and the CIPN trajectory groups using ANOVAs and chi-square tests. Candidate predictors were retained if they were associated at p ≤ 0.25 [50]. Intercorrelations between retained candidate predictors were examined using Pearson’s *r* and Spearman’s rho. In cases of high correlation (≥ 0.7 [56]) between measures, variables with a higher causal priority to CIPN trajectories were considered for inclusion in the regression model [56]. Effect sizes were examined using eta-squared (η²) for continuous variables and Cramer’s V (V) for categorical variables. To determine pre-treatment predictors of CIPN trajectories, multinomial logistic regression was used with criteria for entry and removal set at p ≤ 0.25 and p ≥ 0.05 respectively, with the No CIPN group as the reference category. The level of significance for each covariate and model was set at p ≤ 0.05. Model goodness-of-fit and explained variance were assessed using the Deviance and Pearson chi-square tests and Nagelkerke R². Variance inflation factor (VIF) values were examined to assess potential multicollinearity, with VIF > 10 indicating the presence of multicollinearity [57].

## Results

### Demographic and clinical characteristics

We have previously reported the recruitment and retention flow diagram and reasons for exclusion or withdrawal from T0 to T1 [27]. Briefly, of the 154 participants who initially consented to participate, 148 (96.1%) completed the T0 assessment and 121 (78.6%) completed the T1 assessment. Between T1 and T2, 3 people (1.9%) were no longer interested in participating and withdrew from the study. One hundred and twenty-three people (79.9%) completed the T2 assessment, and two did not due to protocol deviations. This study reports on data from the 102 (66.2%) participants who completed the FACT/GOG-NTX-4 at T0, T1, and T2.

Sociodemographic, cancer, treatment, and other clinical data are presented in Table 1. Participants were aged 51.57 ± 11.24 [range: 24–78] years on average and female. Most (66.7%) received paclitaxel, while 33.3% received docetaxel, and most (67.6%) received treatment with adjuvant intent, while the remaining 32.4% received neoadjuvant treatment. Very few (2.9%) had one or more comorbidity on the CCI. Cancer-related pain burden was high prior to chemotherapy, with most (68.6%) reporting at least one cancer or treatment-related pain condition (e.g., post-surgical pain) at baseline. In addition, 26.5% reported non-cancer-related pain on most days in the past three months.

**Table 1:** Baseline sociodemographic, clinical, and treatment characteristics of participants (n = 102)

| Characteristics | Mean ± SD [range] or n (%) |
| --- | --- |
| **Sociodemographic** |  |
| Age | 51.57 ± 11.24 [24 – 78] |
| Female sex | 102 (100.0) |
| Ethnicity |  |
| Caucasian | 99 (97.1) |
| Other | 3 (2.9) |
| Education |  |
| Elementary/High school | 11 (10.8) |
| Cégep/Professional | 41 (40.2) |
| Bachelor degree | 37 (36.3) |
| Master/doctorate degree | 13 (12.7) |
| Employment status |  |
| Not working due to cancer/treatment | 69 (67.6) |
| Retired | 22 (21.6) |
| Not working due to other reasons | 8 (7.8) |
| Currently working | 3 (2.9) |
| **Cancer and cancer treatment factors** |  |
| Stage |  |
| I | 38 (37.3) |
| II | 39 (38.2) |
| III | 23 (22.5) |
| Missing/Unknown | 2 (2.0) |
| Taxane molecule |  |
| Paclitaxel | 68 (66.7) |
| Docetaxel | 34 (33.3) |
| Adjuvant intent chemotherapy | 69 (67.6) |
| **Clinical data** |  |
| BMI | 25.23 ± 4.29 [17.1 – 39.3] |
| Menopausal stage |  |
| Premenopausal | 44 (43.1) |
| Perimenopausal | 7 (6.9) |
| Postmenopausal | 51 (50.0) |
| CCI ≥ 1 | 3 (2.9) |
| ADS | 0.50 ± 1.03 [0 – 7] |
| Expected pain after chemotherapy | 2.29 ± 1.79 [0 – 8] |
| PHQ |  |
| ≥1 cancer/cancer treatment related pain on most days in the past three months | 70 (68.6) |
| Non-cancer-related pain on most days in the past three months | 27 (26.5) |
| **World Health Organization prescribed analgesic level [97]** |  |
| No analgesic | 12 (11.8) |
| Non-opioid or adjuvant | 72 (70.6) |
| Weak opioid | 5 (4.9) |
| Strong opioid | 12 (11.8) |
| Missing | 1 (0.9) |

Notes. Cégep, *Collège d'enseignement général et professionnel*, an educational institution in Quebec that serves as a bridge between high school and university. Cancer stage was based on pathological staging; if unavailable, clinical staging was used. In case of double breast cancer, the highest stage of the two was retained for the participant. Adjuvant chemotherapy, chemotherapy given after surgery, CCI, Charlson Comorbidity Index, ADS, Anticholinergic Drug Scale, Expected pain after chemotherapy, Average of expected pain ratings obtained immediately and one week post-chemotherapy, PHQ, Pain History Questionnaire, The non-opioid or adjuvant category included acetaminophen, ASA, NSAIDs, and/or adjuvant analgesics (e.g., gabapentin) as the highest prescribed level.

### CIPN trajectory groups

Figure 1 presents mean FACT/GOG-NTX-4 scores at each assessment time point by trajectory group (see also Supplementary Table S2). Based on changes in FACT/GOG-NTX-4 scores from T0, 35 participants (34.3%) were classified in the Persistence group, 26 (25.5%) in the Improvement group, 7 (6.9%) in the Coasting group, and 34 (33.3%) in the No CIPN group. Mean age by CIPN trajectory group was as follows: Persistence, 57.37 ± 8.11 [range: 30–72] years old; Improvement, 46.77 ± 12.67 [range: 28–74] years old; Coasting, 51.43 ± 3.87 [range: 46–57] years old; and No CIPN, 49.20 ± 11.62 [range: 24–78] years old.

**Figure 1:**
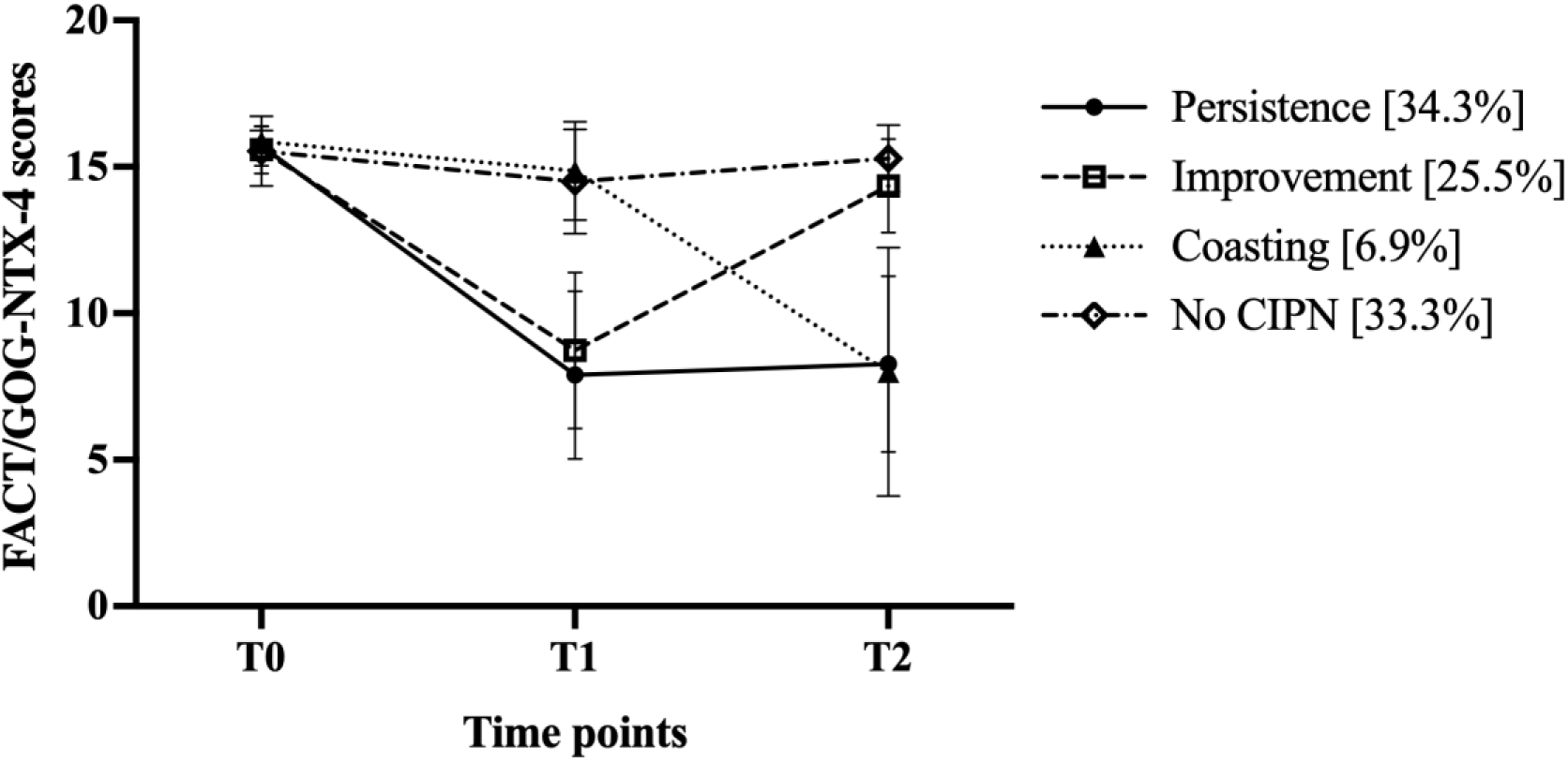
Mean FACT/GOG-NTX-4 scores across trajectory groups. *Notes*. FACT/GOG-NTX-4, Functional Assessment of Cancer Therapy/Gynecologic Oncology Group–Neurotoxicity 4-item scale (0-16). Higher FACT/GOG-NTX-4 scores reflect lower neurotoxicity, T0: prior to the first taxane cycle, T1: end of chemotherapy, at the time of the last taxane infusion (±1 week), T2: three months after the last taxane infusion.

### Bivariate analyses: candidate predictors of CIPN trajectories

Following bivariate analyses (see Supplementary Table S3) between candidate predictors and CIPN trajectory groups (entry threshold p ≤ .25), the following variables were retained and considered for inclusion in the final model: age (p = 0.001), BMI (p = 0.084), foot WDT (p= 0.020), foot VDT (p = 0.137), foot CPT (p = 0.127), hand CDT (p = 0.052), hand CPT (p = 0.102), FACIT-Fatigue (p = 0.004), FACT-PWB (p = 0.001), average expected pain (p = 0.074), and SF-MPQ-2 Neuropathic Pain (p = 0.161). Menopausal stage (premenopausal, perimenopausal, or postmenopausal; p = 0.006) and menopausal status (postmenopausal vs not; p = 0.007) were retained based on chi-square test results. Intercorrelations between retained candidate predictors are shown in Table 2. Examination of intercorrelations revealed that FACIT-Fatigue was strongly correlated with FACT-PWB (*r* = 0.781, p = .01). Given its larger effect size on CIPN trajectories (η² = 0.153 vs. 0.127), FACIT-Fatigue was retained and FACT-PWB excluded. Age was highly correlated with both menopausal stage (ρ = 0.863; p = 0.01) and menopausal status (ρ = 0.812; p = 0.01). Menopausal stage and menopausal status were also strongly associated (ρ = 0.915; p = 0.01). Among these variables, age was retained due to its large effect size (η² = 0.390), compared to medium effect sizes with menopausal stage (V = 0.301) and menopausal status (V = 0.295) [54].

**Table 2:**
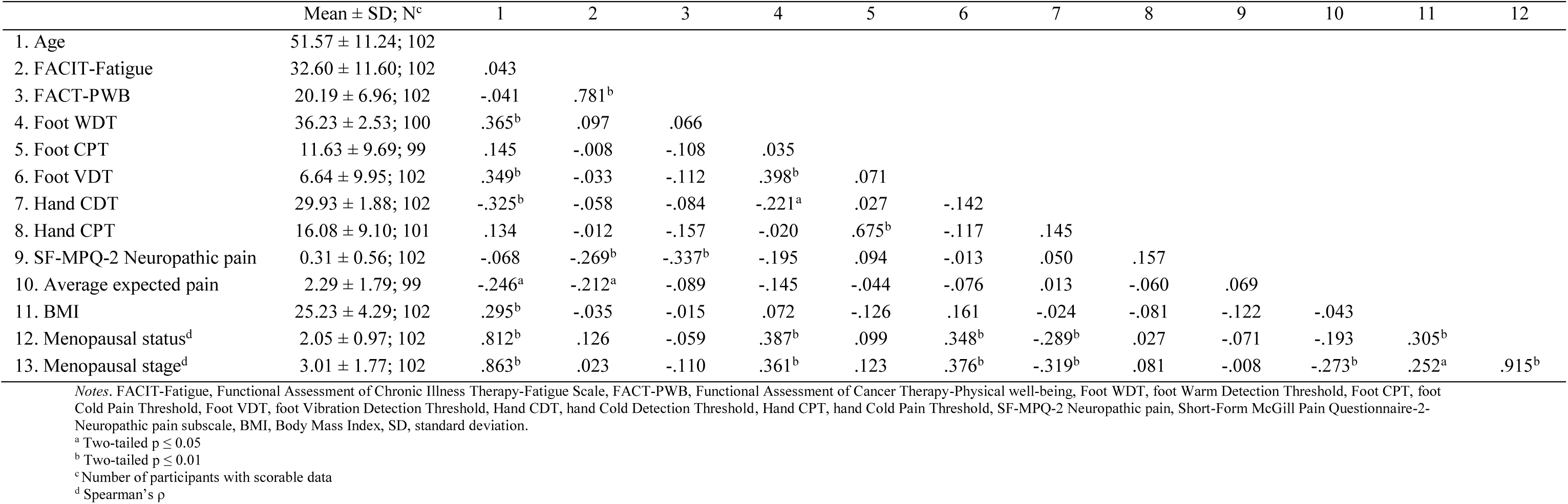
Mean ± SD and correlation matrix of retained T0 candidate predictors.

| | Mean ± SD; N<sup>c</sup> | 1 | 2 | 3 | 4 | 5 | 6 | 7 | 8 | 9 | 10 | 11 | 12 |
| --- | --- | --- | --- | --- | --- | --- | --- | --- | --- | --- | --- | --- | --- |
| 1. Age | 51.57 ± 11.24; 102 | | | | | | | | | | | | |
| 2. FACIT-Fatigue | 32.60 ± 11.60; 102 | .043 | | | | | | | | | | | |
| 3. FACT-PWB | 20.19 ± 6.96; 102 | -.041 | .781<sup>b</sup> | | | | | | | | | | |
| 4. Foot WDT | 36.23 ± 2.53; 100 | .365<sup>b</sup> | .097 | .066 | | | | | | | | | |
| 5. Foot CPT | 11.63 ± 9.69; 99 | .145 | -.008 | -.108 | .035 | | | | | | | | |
| 6. Foot VDT | 6.64 ± 9.95; 102 | .349<sup>b</sup> | -.033 | -.112 | .398<sup>b</sup> | .071 | | | | | | | |
| 7. Hand CDT | 29.93 ± 1.88; 102 | -.325<sup>b</sup> | -.058 | -.084 | -.221<sup>a</sup> | .027 | -.142 | | | | | | |
| 8. Hand CPT | 16.08 ± 9.10; 101 | .134 | -.012 | -.157 | -.020 | .675<sup>b</sup> | -.117 | .145 | | | | | |
| 9. SF-MPQ-2 Neuropathic pain | 0.31 ± 0.56; 102 | -.068 | -.269<sup>b</sup> | -.337<sup>b</sup> | -.195 | .094 | -.013 | .050 | .157 | | | | |
| 10. Average expected pain | 2.29 ± 1.79; 99 | -.246<sup>a</sup> | -.212<sup>a</sup> | -.089 | -.145 | -.044 | -.076 | .013 | -.060 | .069 | | | |
| 11. BMI | 25.23 ± 4.29; 102 | .295<sup>b</sup> | -.035 | -.015 | .072 | -.126 | .161 | -.024 | -.081 | -.122 | -.043 | | |
| 12. Menopausal status<sup>d</sup> | 2.05 ± 0.97; 102 | .812<sup>b</sup> | .126 | -.059 | .387<sup>b</sup> | .099 | .348<sup>b</sup> | -.289<sup>b</sup> | .027 | -.071 | -.193 | .305<sup>b</sup> | |
| 13. Menopausal stage<sup>d</sup> | 3.01 ± 1.77; 102 | .863<sup>b</sup> | .023 | -.110 | .361<sup>b</sup> | .123 | .376<sup>b</sup> | -.319<sup>b</sup> | .081 | -.008 | -.273<sup>b</sup> | .252<sup>a</sup> | .915<sup>b</sup> |

Notes. FACIT-Fatigue, Functional Assessment of Chronic Illness Therapy-Fatigue Scale, FACT-PWB, Functional Assessment of Cancer Therapy-Physical well-being, Foot WDT, foot Warm Detection Threshold, Foot CPT, foot Cold Pain Threshold, Foot VDT, foot Vibration Detection Threshold, Hand CDT, hand Cold Detection Threshold, Hand CPT, hand Cold Pain Threshold, SF-MPQ-2 Neuropathic pain, Short-Form McGill Pain Questionnaire-2-Neuropathic pain subscale, BMI, Body Mass Index, SD, standard deviation.
<sup>a</sup> Two-tailed $p \leq 0.05$
<sup>b</sup> Two-tailed $p \leq 0.01$
<sup>c</sup> Number of participants with scorable data
<sup>d</sup> Spearman's $\rho$

### Multinomial logistic regression: predictors associated with CIPN trajectories

All VIFs were within acceptable limits at ≤ 2.017 [50], suggesting no evidence of problematic multicollinearity among predictors. The model was statistically significant, χ² (30) = 74,805, p < .001, and explained 58.7% of the variance in the outcome categories, as indicated by the Nagelkerke R² value (R² = 0.587). Model goodness-of-fit statistics (Deviance: χ² (255) = 169.749, p = 1.000; Pearson: χ² (255) = 205.740, p = 0.990) were non-significant, indicating an acceptable fit to the observed data.

Table 3 displays multinomial logistic regression results. Compared to the No CIPN group, significant baseline predictors of the Persistence trajectory included older age (OR = 1.120; 95% CI: 1.026 - 1.222), higher average expected pain (OR = 1.630; 95% CI: 1.082 - 2.456) and higher foot CPT (OR = 1.130; 95% CI: 1.018 - 1.254), indicating greater sensitivity. Lower FACIT-Fatigue (OR = 0.904; 95% CI: 0.845 - 0.968), indicating worse fatigue, was the only significant predictor of the Improvement trajectory. No significant predictors were identified for the Coasting trajectory.

**Table 3:** Multinomial logistic regression analysis: retained T0 predictors of CIPN trajectories.

| Predictors <sup>a</sup> | Persistence |  |  |  |  |  |  | Improvement |  |  |  |  |  |  | Coasting |  |  |  |  |  |  |
| --- | --- | --- | --- | --- | --- | --- | --- | --- | --- | --- | --- | --- | --- | --- | --- | --- | --- | --- | --- | --- | --- |
| | B | S.E. | Wald $\chi^2$ | *p* | OR | 95% CI for OR | | B | S.E. | Wald $\chi^2$ | *p* | OR | 95% CI for OR | | B | S.E. | Wald $\chi^2$ | *p* | OR | 95% CI for OR | |
|  |  |  |  |  |  | Lower | Higher |  |  |  |  |  | Lower | Higher |  |  |  |  |  | Lower | Higher |
| Age | **0.113** | **0.045** | **6.394** | **0.011** | **1.120** | **1.026** | **1.222** | -0.033 | 0.036 | 0.841 | 0.359 | 0.967 | 0.902 | 1.038 | -0.040 | 0.062 | 0.410 | 0.522 | 0.961 | 0.852 | 1.085 |
| FACIT-Fatigue | -0.052 | 0.036 | 2.110 | 0.146 | 0.949 | 0.885 | 1.018 | **-0.101** | **0.035** | **8.358** | **0.004** | **0.904** | **0.845** | **0.968** | -0.071 | 0.050 | 2.069 | 0.150 | 0.931 | 0.845 | 1.026 |
| Foot WDT | 0.189 | 0.143 | 1.746 | 0.186 | 1.208 | 0.913 | 1.600 | 0.123 | 0.166 | 0.546 | 0.460 | 1.131 | 0.817 | 1.565 | 0.188 | 0.213 | 0.784 | 0.376 | 1.207 | 0.796 | 1.832 |
| Foot CPT | **0.122** | **0.053** | **5.299** | **0.021** | **1.130** | **1.018** | **1.254** | 0.046 | 0.048 | 0.931 | 0.335 | 1.047 | 0.953 | 1.150 | -0.050 | 0.089 | 0.315 | 0.575 | 0.951 | 0.799 | 1.132 |
| Foot VDT | 0.022 | 0.067 | 0.112 | 0.738 | 1.023 | 0.897 | 1.165 | 0.041 | 0.079 | 0.273 | 0.601 | 1.042 | 0.893 | 1.217 | 0.046 | 0.078 | 0.349 | 0.555 | 1.047 | 0.899 | 1.220 |
| Hand CDT | 0.033 | 0.175 | 0.036 | 0.851 | 1.033 | 0.734 | 1.456 | 0.557 | 0.355 | 2.460 | 0.117 | 1.745 | 0.870 | 3.501 | -0.131 | 0.333 | 0.155 | 0.694 | 0.877 | 0.457 | 1.685 |
| Hand CPT | -0.085 | 0.055 | 2.384 | 0.123 | 0.918 | 0.825 | 1.023 | 0.065 | 0.056 | 1.361 | 0.243 | 1.067 | 0.957 | 1.191 | -0.076 | 0.076 | 0.984 | 0.321 | 0.927 | 0.798 | 1.077 |
| SF-MPQ-2 Neuropathic pain | -1.234 | 0.837 | 2.175 | 0.140 | 0.291 | 0.056 | 1.501 | -0.129 | 0.511 | 0.064 | 0.800 | 0.879 | 0.323 | 2.392 | -2.737 | 1.751 | 2.443 | 0.118 | 0.065 | 0.002 | 2.004 |
| Average expected pain | **0.489** | **0.209** | **5.452** | **0.020** | **1.630** | **1.082** | **2.456** | 0.240 | 0.193 | 1.549 | 0.213 | 1.271 | 0.871 | 1.855 | -0.924 | 0.511 | 3.275 | 0.070 | 0.397 | 0.146 | 1.080 |
| BMI | 0.134 | 0.087 | 2.359 | 0.125 | 1.143 | 0.964 | 1.356 | 0.105 | 0.090 | 1.370 | 0.242 | 1.110 | 0.932 | 1.324 | -0.116 | 0.147 | 0.620 | 0.431 | 0.891 | 0.668 | 1.188 |

Notes. FACIT-Fatigue, Functional Assessment of Chronic Illness Therapy-Fatigue Scale, Foot WDT, foot Warm Detection threshold, Foot CPT, foot Cold Pain Threshold, Foot VDT, foot Vibration Detection Threshold, Hand CDT, hand Cold Detection Threshold, Hand CPT, hand Cold Pain Threshold, SF-MPQ-2 Neuropathic pain, Short-Form McGill Pain Questionnaire-2-Neuropathic pain subscale, BMI, Body Mass Index, CI, Confidence interval, S.E., Standard error, OR, odds ratio.
<sup>a</sup> Reference group: No CIPN

## Discussion

To our knowledge, this study is the first to identify distinct CIPN trajectories and examine how pre-treatment biopsychosocial factors predict distinct CIPN trajectories in people receiving taxane-based chemotherapy for breast cancer. We defined four CIPN trajectories within the first three months following treatment: Persistence, Improvement, Coasting, and No CIPN. Interestingly, there were unique biopsychosocial pre-treatment predictors of the Persistence and Improvement trajectories compared to those with No CIPN. Older age, higher average expected pain after chemotherapy, and greater cold pain sensitivity in the foot were associated with CIPN persistence, whereas lower pre-treatment fatigue predicted the improvement trajectory. These findings extend our understanding of CIPN recovery post-treatment, highlighting heterogeneous trajectories influenced by distinct pre-treatment predictors, as discussed below. Early identification of these vulnerability markers may enhance risk stratification and inform targeted prevention and management strategies to improve QOL.

CIPN trajectories were derived using an anchor-based method based on the MID of the FACT/GOG-NTX-4 scale [37]. Applying this anchor-based MID threshold enhances clinical interpretability by linking score changes to the transition from CTCAE grade 0 to grade 1, reflecting the onset of clinically perceptible CIPN symptoms [37]. In contrast to the previously published CIPN trajectory study [20], which classified trajectories based on severity scores alone, our approach anchors symptom change to a validated, clinically meaningful threshold applied at each time point. This novel use of the MID for trajectory classification enabled the identification of distinct patterns of onset, progression, and improvement. This approach may improve clinical monitoring and supports the use of brief patient-reported measures such as the FACT/GOG-NTX-4, which reduce response burden and enable repeated assessments over time [58].

Using this approach, four distinct CIPN trajectories were identified. The Persistence, Improvement, and No CIPN groups showed comparable prevalence (34.3%, 25.5%, and 33.3%, respectively), whereas Coasting was less frequent (6.9%). Previous studies have reported a wide range of CIPN prevalence across time points and cancer populations. For example, a systematic review and meta-analysis of patients with various cancers treated with different neurotoxic agents reported prevalence rates ranging from 57.7% to 78.4% within the first month following chemotherapy and from 36.4% to 81.6% at three months post-treatment [59]. Similarly, a cohort study of breast cancer patients reported a prevalence of 72.9% at the end of chemotherapy, decreasing to 31.1% by two months post-treatment [60], while estimates among long-term breast cancer survivors range from 11% to over 80% at one to three years post-diagnosis [11]. These wide ranges have been attributed to differences in study design, treatment regimens, and CIPN assessment tools [12, 13]. Our trajectory-based findings suggest that this variability may also reflect underlying heterogeneity in symptom evolution, underscoring the importance of moving beyond single time-point prevalence estimates to identify clinically meaningful subgroups.

The Coasting trajectory, although clinically recognized, remains poorly characterized in longitudinal studies of taxane-induced neuropathy [9, 10]. It has most often been described in association with platinum-based chemotherapeutic agents such as cisplatin and oxaliplatin, which are thought to induce delayed neurotoxicity through sustained dorsal root ganglia damage and accumulation of DNA adducts [9]. Given the limited and inconsistent descriptions of the coasting phenomenon, further research with larger samples is needed to clarify its prevalence and predictors in people receiving taxane-based chemotherapy.

While previous research has identified predictors of CIPN [26, 59, 61, 62], most studies have relied on static assessments at discrete time points, failing to capture the dynamic evolution of symptoms. Although trajectory-based approaches have recently been applied to CIPN to investigate a limited number of sociodemographic, disease-related and other clinical predictors in lymphoma survivors [20], no study has characterized the pre-treatment biopsychosocial predictors of CIPN trajectories in the context of breast cancer. Using this approach, we found that older age predicted membership in the Persistence trajectory but not the Improvement or Coasting, suggesting that age-related neurobiological changes may influence the course of CIPN following taxane-based chemotherapy. This contrasts with trajectory studies of other breast cancer-related symptoms, where younger age has been linked to persistent post-surgical pain [19] and depressive symptoms [17], likely reflecting distinct neurotoxic mechanisms in CIPN. In support of this, our findings are consistent with the only other CIPN trajectory study, where older age was associated with membership in a high CIPN group characterized by symptom escalation during chemotherapy and persistence up to ten weeks post-treatment among people treated with vincristine for lymphoma [20].

Aging induces degenerative changes in the peripheral nervous system [63, 64], including reduced nerve conduction velocity, loss of myelinated and unmyelinated fibers, myelin fragmentation, and impaired axonal regeneration and reinnervation [65–67], potentially heightening susceptibility to taxane-induced neurotoxicity. Evidence from diabetic populations further demonstrates that aging amplifies vulnerability to peripheral nerve damage [68]. Aging is also associated with a decline in endogenous pain inhibition and enhanced central sensitization [69, 70], which may further impair recovery from nerve injury. Consistent with findings of this study, a longitudinal study of CIPN in gynecologic cancer patients demonstrated that, while CIPN increased comparably in older and younger adults during chemotherapy, only younger adults demonstrated post-treatment recovery whereas CIPN persisted in older adults [71]. Taken together, these results suggest that age-related structural and functional deficits in both the peripheral and central nervous systems may contribute to CIPN persistence among older people.

Higher foot cold pain sensitivity (i.e., higher CPT) predicted membership in the Persistence, but not Improvement or Coasting trajectories, suggesting that altered nociceptive processing prior to chemotherapy may predispose patients to persistent CIPN in the first three months after chemotherapy. This is consistent with evidence showing that cold hyperalgesia is a common feature of neuropathic pain [72] and may reflect central sensitization and impaired pain modulation [73]. In the context of CIPN specifically, cold hyperalgesia during oxaliplatin treatment predicted the development of severe neuropathy [74].

While previous work has underscored the prognostic value of cold sensitivity, it has been limited to intra-treatment assessments or global CIPN severity rather than to longitudinal symptom trajectories [74]. Baseline QST measures have demonstrated prognostic value across modalities. For example, lower baseline vibration detection threshold has predicted clinically significant neuropathy six months after oxaliplatin [75], and higher baseline warm detection thresholds (i.e., indicating reduced thermal sensitivity) have predicted grade 2 or 3 neuropathy one year after oxaliplatin [76]. Our findings extend this work by demonstrating that pre-treatment cold pain sensitivity predicts vulnerability to a Persistent CIPN trajectory, not merely higher overall severity, highlighting the potential value of baseline QST for early risk stratification and tailored monitoring strategies. Notably, although aging is associated with decreased cold pain sensitivity in healthy adults [77], the predictive role of cold pain hyperalgesia in the present study suggests that pre-existing nociceptive sensitization, rather than age-related hyposensitivity, may represent a distinct vulnerability factor for CIPN persistence. Future studies should examine whether age moderates the relationship between baseline cold pain sensitivity and CIPN trajectories.

Intriguingly, greater pre-treatment expectations of pain after chemotherapy predicted membership in the Persistence, but not Improvement or Coasting trajectories, suggesting that pre-treatment pain expectations may influence CIPN chronicity in the first three months after chemotherapy. Notably, this effect was specific to pain expectations rather than pre-treatment self-reported pain intensity or qualities, which was not associated with trajectory membership, suggesting that the anticipation of future pain may be a more important predictor of CIPN persistence than baseline pain experience itself. This finding is consistent with research in chronic non-cancer pain that has shown that negative pain expectations predict greater long-term pain severity and functional disability [78, 79]. Our measure of CIPN assessed tingling and numbness rather than painful symptoms specifically, consistent with evidence that CIPN can often manifest as non-painful paresthesia [80] characterized by abnormal sensory experiences such as tingling, numbness, or hyposensitivity. These sensations may be strongly shaped by cognitive and emotional processes, including attentional focus and maladaptive interpretations of bodily sensations [81, 82]. Heightened pain expectations may reflect hypervigilance to bodily sensations and negative interpretation biases (i.e., a tendency to perceive or interpret bodily sensations as threatening or indicative of harm), amplifying the salience of minor or ambiguous sensory changes [83, 84]. Consistent with this, a recent study in breast and gynaecological cancer survivors found that fear of cancer recurrence, a related form of threat appraisal prospectively predicted future pain, whereas pain did not predict future fear, suggesting that negative cognitive appraisals may contribute to symptom persistence over time [85]. These processes may further limit engagement in adaptive behaviors, reinforcing symptom chronicity over time [86].

Neuroimaging research has shown that the expectation of pain enhances activity in regions involved in affective and sensory processing, including the anterior cingulate cortex and the parietal operculum/posterior insula [87]. These regions are part of broader brain networks also implicated in the subjective experience and modulation of somatosensory input, suggesting that heightened pain expectations may prime or sensitize central pathways, potentially increasing the perceived intensity before any nociceptive input occurs [88–90]. Taken together, these findings suggest that heightened pre-treatment pain expectations may increase attentional focus toward sensory input, which in turn could intensify and prolong CIPN symptoms. Pre-treatment expectations may therefore represent a modifiable risk factor, supporting the implementation of targeted educational and psychotherapeutic strategies.

The only predictor of the Improvement trajectory was pre-treatment fatigue, where lower scores on the FACIT-Fatigue scale, indicating greater fatigue, predicted membership in the improvement trajectory, but not Persistence or Coasting. This is consistent with prior research identifying fatigue as a pre-treatment risk factor for CIPN. In a longitudinal study of women with breast cancer receiving taxane- or platinum-based chemotherapy, higher levels of fatigue, anxiety, and depression prior to treatment were among the strongest predictors of neuropathic symptoms six weeks after chemotherapy [26]. Fatigue has also been linked to biological mechanisms implicated in CIPN, including neuroinflammation and mitochondrial dysfunction [91–93]. More precisely, cancer-related fatigue has been associated with low-grade systemic inflammation and immune-mediated alterations affecting both central and peripheral nervous systems [94, 95]. Our results expand of these findings by showing that fatigue does not uniformly increase vulnerability to CIPN but may also be linked to subsequent improvement. This trajectory-specific association may suggest that high pre-treatment fatigue may act as a risk factor for post-treatment transient neural sensitization rather than irreversible injury following exposure to neurotoxic agents. Fatigue may be a modifiable risk factor [96], influencing not only vulnerability to symptom onset but also the likelihood of subsequent resolution, thereby underscoring its relevance for both prediction and intervention. Further research is needed to examine whether early fatigue interventions can influence CIPN symptom trajectories.

Several limitations should be considered in the interpretation of the present findings. First, the relatively small sample size limited statistical power and may have constrained the identification of a broader range of biopsychosocial predictors, particularly for the Coasting trajectory, which was underrepresented. Larger samples are needed for more robust trajectory modeling. Second, the sample was predominantly composed of well-educated white participants with few comorbidities, which may limit the generalizability of the findings to more diverse populations. Third, the MID [37] used to define clinically meaningful changes in CIPN as measured by the FACT/GOG-NTX-4 may not fully capture individual variability in CIPN experience, as this outcome primarily captures sensory symptoms rather than the painful dimension of CIPN. Future trajectory research should use a more comprehensive, multidimensional measure of the CIPN experience to better characterize CIPN trajectories. The study was also limited to two post-treatment time points, reducing our ability to capture short-term fluctuations and longer-term symptom evolution. Incorporating intra-treatment assessments and longer post-treatment follow-ups would provide a more comprehensive understanding of CIPN trajectories. Additionally, given the exploratory nature of this study and the number of variables tested, the familywise error rate was not controlled for, and findings should be interpreted with caution. Participants were all treated for early-stage breast cancer. CIPN-related processes may differ in the context of advanced disease, where treatment goals and underlying vulnerabilities may vary. Finally, excluding participants with comorbidities potentially associated with peripheral neuropathy may have restricted the range of CIPN trajectories and associated biopsychosocial predictors identified. Future studies should include participants with these comorbidities to better characterize the full spectrum of CIPN trajectories and their predictors.

## Conclusion

By identifying distinct CIPN trajectories, this study highlights the limitations of relying on single time-point prevalence estimates to understand CIPN and demonstrates that the wide variability in CIPN prevalence reported in previous research may reflect underlying differences in symptom courses in addition to cross-study methodological variability. This framework also identifies pre-treatment biopsychosocial predictors of different symptom pathways, highlighting the potential for a more nuanced approach to understanding risk identification for prolonged post-treatment CIPN. A better understanding of elevated risk of persistent CIPN could inform targeted supportive care efforts, such as enhanced symptom monitoring and early, preventative interventions.

## Statements and Declarations

### Author Contributions

Conceptualization: Charles-Antoine Auger, Lynn R. Gauthier, Antoine Frasie, Lucia Gagliese

Methodology: Charles-Antoine Auger, Lynn R. Gauthier, Antoine Frasie, Anne Dionne, Robert H. Dworkin, Lucia Gagliese, Jennifer S. Gewandter, Philip L. Jackson, Sophie Lauzier, Julie Lemieux, Josée Savard

Formal analysis and investigation: Charles-Antoine Auger, Lynn R. Gauthier, Antoine Frasie

Writing – original draft preparation: All authors Financial support: Lynn R. Gauthier

Writing – review and editing: All authors Funding acquisition: Lynn R. Gauthier

Resources: Lynn R. Gauthier, Julie Lemieux, Philip L. Jackson, Anne Dionne, Maud

Bouffard, Frédérique Therrien, Sarah Béland

Supervision: Lynn R. Gauthier

### Ethics approval

Approval was received from the Research Ethics Committee of the CHU de Québec-Université Laval (REB#2017-3312).

### Prior presentation

Part of these results were presented as a poster at the 2025 Canadian Cancer Research Conference.

### Consent to participate

Written informed consent to participate was obtained from all study participants.

### Consent to publish

Not applicable.

### Data availability

Data and materials may be made available upon request.

### Supplementary Information

The online version contains supplementary material.

## Acknowledgments

We would like to thank all study participants for their time and contributions, as well as the staff at the Centre des maladies du sein du Québec.

