## Supplementary Material for "Pre-treatment biopsychosocial predictors of chemotherapy-induced peripheral neuropathy trajectories in people with breast cancer"

### Supplementary Table S1

Strengthening the reporting of observational studies in epidemiology (STROBE) guidelines for cohort studies checklist

| Item No | Recommendation | Page No. | Reported (Yes/No/N/A) |
|---|---|---|---|
| **Title and abstract** | | | |
| 1(a) | Indicate the study's design with a commonly used term in the title or the abstract | 3 (abstract) | Yes |
| 1(b) | Provide in the abstract an informative and balanced summary of what was done and what was found | 3 | Yes |
| **Introduction** | | | |
| 2 | Explain the scientific background and rationale for the investigation being reported | 5-7 (intro) | Yes |
| 3 | State specific objectives, including any prespecified hypotheses | 7 | Yes |
| **Methods** | | | |
| 4 | Present key elements of study design early in the paper | 7 (methods) | Yes |
| 5 | Describe the setting, locations, and relevant dates, including periods of recruitment, exposure, follow-up, and data collection | 7, 8 | Yes |
| 6(a) | Give the eligibility criteria, and the sources and methods of selection of participants. Describe methods of follow-up | 7, 8 | Yes |
| 6(b) | For matched studies, give matching criteria and number of exposed and unexposed | N/A | N/A |
| 7 | Clearly define all outcomes, exposures, predictors, potential confounders, and effect modifiers. Give diagnostic criteria, if applicable | 8-14 | Yes |
| 8 | For each variable of interest, give sources of data and details of methods of assessment (measurement). Describe comparability of assessment methods if there is more than one group | 8-13 | Yes |
| 9 | Describe any efforts to address potential sources of bias | 7, 8 | Yes |
| 10 | Explain how the study size was arrived at | 13 | Yes |
| 11 | Explain how quantitative variables were handled in the analyses. If applicable, describe which groupings were chosen and why | 13, 14 | Yes |
| 12(a) | Describe all statistical methods, including those used to control for confounding | 13, 14 | Yes |
| 12(b) | Describe any methods used to examine subgroups and interactions | N/A | N/A |
| 12(c) | Explain how missing data were addressed | 13 | Yes |
| 12(d) | If applicable, explain how loss to follow-up was addressed | N/A | N/A |
| 12(e) | Describe any sensitivity analyses | N/A | No |
| **Results** | | | |
| 13(a) | Report numbers of individuals at each stage of study — e.g., numbers potentially eligible, examined for eligibility, confirmed eligible, included in the study, completing follow-up, and analysed | 14 | Yes |
| 13(b) | Give reasons for non-participation at each stage | 14 | Yes |
| 13(c) | Consider use of a flow diagram | 14 | Yes |
| 14(a) | Give characteristics of study participants (e.g., demographic, clinical, social) and information on exposures and potential confounders | 14, 15, Table 1 | Yes |
| 14(b) | Indicate number of participants with missing data for each variable of interest | Table 1 | Yes |
| 14(c) | Summarise follow-up time (e.g., average and total amount) | 8, 14, 15 | Yes |
| 15 | Report numbers of outcome events or summary measures over time | 15, 16, Figure 1, Table 2, 3, Table S1, S2 | Yes |
| 16(a) | Give unadjusted estimates and, if applicable, confounder-adjusted estimates and their precision (e.g., 95% confidence interval). Make clear which confounders were adjusted for and why they were included | N/A | Yes |
| 16(b) | Report category boundaries when continuous variables were categorized | 9, 10 | Yes |

| | | | |
|---|---|---|---|
| 16(c) | If relevant, consider translating estimates of relative risk into absolute risk for a meaningful time period | N/A | N/A |
| 17 | Report other analyses done — e.g., analyses of subgroups and interactions, and sensitivity analyses | 13 (power analysis) | Yes |
| **Discussion** | | | |
| 18 | Summarise key results with reference to study objectives | 17 | Yes |
| 19 | Discuss limitations of the study, taking into account sources of potential bias or imprecision. Discuss both direction and magnitude of any potential bias | 23 | Yes |
| 20 | Give a cautious overall interpretation of results considering objectives, limitations, multiplicity of analyses, results from similar studies, and other relevant evidence | 17-23 | Yes |
| 21 | Discuss the generalisability (external validity) of the study results | 17-24 | Yes |
| **Other information** | | | |
| 22 | Give the source of funding and the role of the funders for the present study and, if applicable, for the original study on which the present article is based | See Statements and Declarations | Yes |

*Note*: N/A indicates items not applicable to this study design or analysis.

**Supplementary Table S2**

FACT/GOG-NTX-4 scores across assessment time points by CIPN trajectory group

| CIPN trajectory groups | N (%) | Time points (mean ± SD) | | |
|---|---|---|---|---|
| | | **T0** | **T1** | **T2** |
| Persistence | 35 (34.3) | 15.71 ± 0.67 | 7.89 ± 2.86 | 8.27 ± 3.00 |
| Improvement | 26 (25.5) | 15.58 ± 0.81 | 8.73 ± 2.66 | 14.35 ± 1.60 |
| Coasting | 7 (6.9) | 15.86 ± 0.38 | 14.86 ± 1.68 | 8.00 ± 4.24 |
| No MID-CIPN | 34 (33.3) | 15.53 ± 1.19 | 14.50 ± 1.78 | 15.29 ± 1.14 |
| Total | 102 (100.0) | 15.63 ± 0.89 | 10.78 ± 3.93 | 12.14 ± 4.02 |

*Notes.* FACT/GOG-NTX-4, Functional Assessment of Cancer Therapy/Gynecologic Oncology Group–Neurotoxicity 4-item scale (0-16). Higher FACT/GOG-NTX-4 scores reflect lower neurotoxicity.

**Supplementary Table S3**

Results of bivariate analyses between candidate predictors and CIPN trajectory groups

| Variable | Statistic | df | *p* | Effect size | Decision |
|---|---|---|---|---|---|
| **Continuous variables (ANOVA)** | | | | | |
| Age | F = 5.909 | 3, 98 | .001 | $\eta^2$ = .390 | Retained |
| FACIT-Fatigue | F = 4.751 | 3, 98 | .004 | $\eta^2$ = .153 | Retained |
| FACT-PWB | F = 5.910 | 3, 98 | .001 | $\eta^2$ = .127 | Excluded (high intercorrelation)[a] |
| Foot CDT | F = 0.716 | 3, 98 | .545 | $\eta^2$ = .021 | Excluded (p > .25) |
| Foot WDT | F = 3.428 | 3, 96 | .020 | $\eta^2$ = .097 | Retained |
| Foot CPT | F = 1.946 | 3, 95 | .127 | $\eta^2$ = .058 | Retained |
| Foot HPT | F = 0.799 | 3, 95 | .497 | $\eta^2$ = .025 | Excluded (p > .25) |
| Foot VDT | F = 1.887 | 3, 98 | .137 | $\eta^2$ = .055 | Retained |
| Foot MDT | F = 0.233 | 3, 97 | .873 | $\eta^2$ = .007 | Excluded (p > .25) |
| Hand CDT | F = 2.668 | 3, 98 | .052 | $\eta^2$ = .076 | Retained |
| Hand WDT | F = 1.116 | 3, 98 | .347 | $\eta^2$ = .033 | Excluded (p > .25) |
| Hand CPT | F = 2.127 | 3, 97 | .102 | $\eta^2$ = .062 | Retained |
| Hand HPT | F = 0.939 | 3, 96 | .425 | $\eta^2$ = .029 | Excluded (p > .25) |
| Hand VDT | F = 1.164 | 3, 97 | .327 | $\eta^2$ = .035 | Excluded (p > .25) |
| Hand MDT | F = 0.822 | 3, 98 | .485 | $\eta^2$ = .025 | Excluded (p > .25) |
| SF-MPQ-2 Neuropathic pain | F = 1.753 | 3, 98 | .161 | $\eta^2$ = .051 | Retained |
| Average expected pain | F = 2.384 | 3, 95 | .074 | $\eta^2$ = .070 | Retained |
| BMI | F = 2.278 | 3, 98 | .084 | $\eta^2$ = .065 | Retained |
| TUG | F = 0.670 | 3, 97 | .572 | $\eta^2$ = .020 | Excluded (p > .25) |
| PSQI | F = 0.247 | 3, 98 | .863 | $\eta^2$ = .008 | Excluded (p > .25) |
| CES-D | F = 0.810 | 3, 98 | .491 | $\eta^2$ = .024 | Excluded (p > .25) |
| PCS | F = 0.848 | 3, 97 | .471 | $\eta^2$ = .026 | Excluded (p > .25) |
| FACT-FWB | F = 0.732 | 3, 98 | .536 | $\eta^2$ = .022 | Excluded (p > .25) |
| FACT-SWB | F = 0.328 | 3, 98 | .805 | $\eta^2$ = .010 | Excluded (p > .25) |
| FACT-EWB | F = 0.220 | 3, 98 | .882 | $\eta^2$ = .007 | Excluded (p > .25) |
| BPI pain intensity | F = 1.333 | 3, 93 | .269 | $\eta^2$ = .041 | Excluded (p > .25) |
| BPI pain interference | F = 1.312 | 3, 97 | .275 | $\eta^2$ = .039 | Excluded (p > .25) |
| NPSI | F = 1.315 | 3, 98 | .274 | $\eta^2$ = .039 | Excluded (p > .25) |
| FACT-Taxane | F = 1.038 | 3, 98 | .379 | $\eta^2$ = .031 | Excluded (p > .25) |
| SF-MPQ-2 Total | F = 0.530 | 3, 96 | .663 | $\eta^2$ = .016 | Excluded (p > .25) |
| SF-MPQ-2 Continuous pain | F = 0.296 | 3, 98 | .828 | $\eta^2$ = .009 | Excluded (p > .25) |
| SF-MPQ-2 Affective pain | F = 0.594 | 3, 98 | .621 | $\eta^2$ = .018 | Excluded (p > .25) |
| KPS score | F = 0.990 | 3, 98 | .401 | $\eta^2$ = .029 | Excluded (p > .25) |
| **Categorical variables (chi-square)** | | | | | |
| Menopausal status | $\chi^2$ = 17.743 | 6 | .007 | V = .295 | Excluded (high intercorrelation)[b] |
| Menopausal stage | $\chi^2$ = 27.710 | 12 | .006 | V = .301 | Excluded (high intercorrelation)[b] |
| Charlson Comorbidity Index | $\chi^2$ = 3.299 | 3 | .348 | V = .180 | Excluded (p > .25) |
| Falls/near-falls (past week) | $\chi^2$ = 2.022 | 3 | .568 | V = .146 | Excluded (p > .25) |
| SPPB | $\chi^2$ = 1.580 | 3 | .664 | V = .124 | Excluded (p > .25) |
| Cancer-related pain (PHQ) | $\chi^2$ = 3.855 | 3 | .278 | V = .194 | Excluded (p > .25) |
| Non-cancer pain (PHQ) | $\chi^2$ = 1.652 | 3 | .648 | V = .127 | Excluded (p > .25) |
| Acupuncture use | $\chi^2$ = 1.286 | 3 | .733 | V = .112 | Excluded (p > .25) |
| Opioid prescription | $\chi^2$ = 2.486 | 3 | .478 | V = .156 | Excluded (p > .25) |
| Antidepressant prescription | — | — | — | — | Excluded[c] |
| Gabapentin/pregabalin prescription | $\chi^2$ = 0.962 | 3 | .810 | V = .097 | Excluded (p > .25) |
| TENS/Scrambler therapy | $\chi^2$ = 2.952 | 3 | .399 | V = .170 | Excluded (p > .25) |
| Exercise (past week) | $\chi^2$ = 7.117 | 3 | .068 | V = .264 | Excluded (p > .25) |
| ADS score | $\chi^2$ = 14.409 | 12 | .275 | V = .217 | Excluded (p > .25) |
| Caucasian ethnicity | $\chi^2$ = 2.031 | 3 | .566 | V = .141 | Excluded (p > .25) |
| Religion | $\chi^2$ = 7.450 | 6 | .281 | V = .191 | Excluded (p > .25) |
| Marital status | $\chi^2$ = 0.739 | 3 | .864 | V = .085 | Excluded (p > .25) |
| Living arrangement | $\chi^2$ = 1.447 | 3 | .695 | V = .119 | Excluded (p > .25) |
| Walking aid use | $\chi^2$ = 3.853 | 3 | .278 | V = .194 | Excluded (p > .25) |
| Education level | $\chi^2$ = 7.144 | 9 | .622 | V = .153 | Excluded (p > .25) |

***Notes***. FACIT-Fatigue, Functional Assessment of Chronic Illness Therapy-Fatigue Scale; FACT-PWB, Functional Assessment of Cancer Therapy-Physical Well-Being; FACT-FWB, Functional Assessment of Cancer Therapy-Functional Well-Being; FACT-SWB, Functional Assessment of Cancer Therapy-Social/Family Well-Being; FACT-EWB, Functional Assessment of Cancer Therapy-Emotional Well-Being; WDT, warm detection threshold; CPT, cold pain threshold; CDT, cold detection threshold; VDT, vibration detection threshold; HPT, heat pain threshold; MDT, mechanical detection threshold; SF-MPQ-2, Short-Form McGill Pain Questionnaire-2; BMI, body mass index; TUG, Timed Up and Go; PSQI, Pittsburgh Sleep Quality Index; CES-D, Center for Epidemiologic Studies Depression Scale; PCS, Pain Catastrophizing Scale; BPI, Brief Pain Inventory; NPSI, Neuropathic Pain Symptom Inventory; KPS, Karnofsky Performance Status; PHQ, Pain History Questionnaire; SPPB, Short Physical Performance Battery; ADS, Anticholinergic Drug Scale; TENS, transcutaneous electrical nerve stimulation; V, Cramer's V; $\eta^2$, eta-squared.

[a] FACT-PWB was excluded due to high intercorrelation with FACIT-Fatigue ($r = 0.781$, $p < .01$). FACIT-Fatigue was retained based on its larger effect size ($\eta^2 = .153$ vs. $\eta^2 = .127$).

[b] Menopausal status and menopausal stage were excluded due to high intercorrelations with age ($\rho = 0.812$ and $\rho = 0.863$, respectively, both $p < .01$) and with each other ($\rho = 0.915$, $p < .01$). Age was retained based on its larger effect size ($\eta^2 = .390$).

[c] Antidepressant prescription was not analyzed because no participant reported antidepressant use at baseline.
